# Ergothioneine Supplementation and Patient-Reported Urological Symptoms in Adults with Mildly Reduced or Borderline Renal Function: An Exploratory Open-Label Trial

**DOI:** 10.64898/2026.07.08.26356008

**Authors:** Fan Rong, Zhili Wu, Yuanyuan Xu, Wei Liu, Guohan Zhou, Wei Ding, Juan Cao, Guohua Xiao, Dalin Xu, Huan Zhou

## Abstract

**Background:** Early renal function decline is often accompanied by bothersome urological symptoms, yet effective early-stage nutritional interventions remain limited. L-Ergothioneine (EGT), a diet-derived antioxidant concentrated in renal tissue via the OCTN1 transporter, has shown renoprotective potential preclinically, but human interventional data are sparse.

**Methods:** In this single-center, open-label, self-controlled trial, 31 adults (aged 45–70 years) with early renal function decline and persistent urological symptoms (≥3 months) received oral EGT (120 mg/day) for 90 days; 27 completed the study. Participants served as their own controls. The primary outcome was the within-subject change in eGFR (CKD-EPI 2021 creatinine); secondary outcomes included cystatin C-based eGFR, serum creatinine, UACR, a 10-item voiding diary, and a low-back-pain visual analogue scale (VAS). Within-subject changes were assessed by paired t-test or Wilcoxon signed-rank test.

**Results:** Creatinine-based eGFR increased from 86.04 ± 17.89 to 93.25 ± 19.00 mL/min/1.73 m² (+8.4%; p = 0.0016) and serum creatinine fell by 7.0% (p = 0.015). However, cystatin C-based eGFR and serum cystatin C were unchanged (p = 0.31 and p = 0.99), so the filtration signal was not corroborated by an independent, muscle-mass-independent marker. UACR showed a non-significant downward trend. Patient-reported outcomes improved most robustly: the total voiding diary score decreased by 57.2% (p < 0.0001) and low-back-pain VAS by 67.2% (p = 0.0002), with significant relief of urgency, frequency, and voiding difficulty. No product-related adverse events occurred.

**Conclusions:** In this uncontrolled study, 90-day EGT supplementation was associated with marked improvement in urological symptoms and in creatinine-based eGFR, although the latter was not confirmed by cystatin C. These changes cannot be attributed to EGT alone and may substantially reflect placebo and natural-history effects. The findings are hypothesis-generating and warrant confirmation in a randomized, placebo-controlled trial using validated symptom instruments.

**Trial Registration:** ChiCTR2500108897; Prospectively registered on 2025-09-08.

## 1. Introduction

Chronic kidney disease (CKD) has emerged as a major global public health priority, affecting approximately 10–12% of adults worldwide and imposing a substantial and growing burden on health systems [1,2]. The natural history of CKD is characterized by a slow, often clinically silent attrition of functional nephrons, punctuated only late by overt symptoms [3]. The early window of renal functional decline — when the estimated glomerular filtration rate (eGFR) begins to drift downward but before irreversible structural damage accrues — is therefore widely regarded as the most opportune period for intervention. Yet, paradoxically, it is also the period for which the fewest evidence-based therapeutic options exist; current management is dominated by blood-pressure and glycemic control and by renin-angiotensin system blockade, strategies aimed largely at slowing progression rather than actively supporting renal function or relieving symptoms [3,6].

Compounding this therapeutic gap, individuals in the early stages of renal decline frequently report a constellation of urological symptoms — urinary frequency and urgency, foamy or darkened urine, nocturia, and low-back soreness — that are seldom captured by conventional biochemical monitoring yet substantially impair quality of life [4,5]. These patient-reported outcomes (PROs) have gained increasing recognition from regulatory and nephrology bodies as essential complements to laboratory endpoints, because they reflect the dimensions of health that patients themselves value most [4]. An intervention that could simultaneously support objective filtration and relieve these subjective symptoms would address a genuine unmet need.

Oxidative stress and mitochondrial compromise are well-documented contributors to renal injury [7]. The kidney is a highly metabolic, mitochondria-rich organ whose proximal tubular cells are particularly vulnerable to reactive oxygen species (ROS); ROS accumulation disrupts the glomerular filtration barrier, promotes tubular cell senescence, and drives the progression from acute injury to fibrosis [6,7]. This pathophysiology has motivated numerous antioxidant trials, yet conventional systemic antioxidants such as vitamins C and E have largely failed to show consistent renoprotection [7]. This translational failure is widely attributed to their lack of tissue-specific targeting: they cannot achieve and sustain therapeutic concentrations within the renal mitochondrial compartment where oxidative injury originates [10,11].

L-Ergothioneine (EGT), a naturally occurring sulfur-containing amino acid derived primarily from dietary fungi and obtainable only through the diet in humans, is mechanistically distinct from these failed agents [8,9]. Its tissue distribution is not governed by passive diffusion but by a specific, high-affinity transporter — the organic cation transporter novel type 1 (OCTN1, encoded by SLC22A4) [8,12]. OCTN1 is highly expressed on the apical membrane of renal proximal tubular cells and within mitochondrial membranes, enabling active reabsorption and preferential accumulation of EGT in kidney tissue [8,11]. This transporter-mediated targeting allows EGT to reach the very compartment where conventional antioxidants fail. Preclinically, EGT mitigates cisplatin-induced nephrotoxicity through Nrf2 activation and NF-κB suppression [13], stabilizes mitochondrial function, and attenuates the transition from acute kidney injury to chronic kidney disease [15,18]; it also prevents cystine lithiasis in a murine model of cystinuria [14]. Notably, EGT is markedly depleted in the erythrocytes and plasma of patients receiving hemodialysis, and EGT loss has been proposed as a feature of advanced renal failure, suggesting that maintaining EGT homeostasis may be relevant to renal resilience [16].

Despite this compelling preclinical rationale, human interventional evidence for EGT in renal health remains confined largely to observational or preclinical work [13–15]; no clinical study has, to our knowledge, evaluated EGT supplementation against both objective filtration markers and patient-reported urological symptoms. As an initial step toward clinical evaluation, we conducted a 90-day, single-center, open-label, self-controlled trial in adults with early renal functional decline. Recognizing that an uncontrolled design cannot establish efficacy, the study was deliberately framed as a proof-of-concept investigation intended to generate effect-size estimates, characterize the direction and magnitude of change across complementary objective and subjective endpoints, and inform the design of a future randomized controlled trial.

## 2. Materials and Methods

### 2.1 Study Design and Ethics

A single-center, open-label, self-controlled (within-subject, before-after) clinical trial conducted at The First Affiliated Hospital of Anhui Medical University. The protocol was approved by the Clinical Medical Research Ethics Committee of the First Affiliated Hospital of Anhui Medical University (Approval No. PJ2025-10-14(KY)) and conducted in accordance with the Declaration of Helsinki and GCP guidelines. All participants provided written informed consent. Trial period: 2025-09-28 to 2026-03-13.

### 2.2 Participants

Inclusion: (1) age 45–70 years, either sex; (2) early renal function decline, defined by meeting any one of: (a) 45 ≤ eGFR < 59 with UACR ≤ 30 mg/g; (b) 60 ≤ eGFR ≤ 89 with UACR ≤ 300 mg/g; (c) eGFR ≥ 90 with 30 ≤ UACR ≤ 300 mg/g; (3) urological symptoms (urinary frequency/urgency, darkened urine, foamy urine, low-back soreness) persisting ≥ 3 months; (4) written informed consent.

Exclusion: abnormal liver function (AST/ALT/ALP > 3× ULN) or active hepatitis/ cirrhosis; poorly controlled hypertension/hyperlipidemia/hyperglycemia/hyperuricemia; high-risk CKD; active nephritis with persistent hematuria/oliguria/edema; immunosuppressant use for renal disease; recurrent urinary/genital infection; recent acute cardiac events or stroke; psychiatric or autoimmune disease history; GI disease affecting drug absorption; participation in other trials within 3 months; pregnancy/lactation/pregnancy plans within 6 months; or investigator judgment.

Enrollment: 85 screened; 31 enrolled; 27 completed the 90-day intervention (4 discontinued for personal reasons). The safety population comprised all 31 enrolled participants; the efficacy analysis comprised the 27 completers.

### 2.3 Intervention

Oral EGT capsules (30 mg/capsule; Jiangsu Gene III Biotechnology Co., Ltd.) at 120 mg/day (2 capsules twice daily) for 90 consecutive days. Compliance was assessed by pill count at each visit.

### 2.4 Outcomes

Primary: Efficacy (D0, D60, D90): eGFR (CKD-EPI 2021 creatinine); UACR; serum creatinine, urea, UREA/CREA, uric acid, cystatin C; urinary electrolytes (Na, K, Cl). Voiding diary (D0, D30, D60, D90): a study-specific 10-item voiding symptom diary plus a low-back-pain VAS (see below).

Exploratory marker (D0, D60, D90): urinary 8-hydroxy-2’-deoxyguanosine (8-OHdG), a marker of systemic oxidative DNA damage, measured by high-sensitivity ELISA and normalized to urinary creatinine (ng/mg Cr).

Safety (D0, D60, D90): complete blood count, liver/renal function, urinalysis, urine pregnancy test (women); adverse event monitoring.

Voiding diary instrument. Urological symptoms were recorded using a study-specific 10-item diary covering: (Q1) sensation of incomplete emptying, (Q2) urinary frequency (voiding interval < 2 h), (Q3) intermittency, (Q4) urgency, (Q5) weak stream, (Q6) straining to void, (Q7) darkened urine, (Q8) increased urinary foam, (Q9) nocturia, and (Q10) low-back soreness. Each item was scored 0–5 according to symptom frequency (total score range 0–50; higher scores indicate more severe symptoms). Low-back pain was additionally rated on a 0–10 cm visual analogue scale (VAS). This instrument was developed for the present study and has not undergone formal psychometric validation; symptom findings should therefore be interpreted as exploratory.

### 2.5 Statistical Analysis

Analyses used SAS 9.4. Continuous variables are presented as mean ± SD. Within-subject changes from baseline to each follow-up were assessed by paired t-test for normally distributed data, or Wilcoxon signed-rank test otherwise. Significance was set at α = 0.05 (two-sided). An exploratory subgroup analysis compared eGFR change between participants with baseline UACR ≤ 30 vs > 30 mg/g. As a single-arm study, no between-group comparison was performed.

## 3. Results

### 3.1 Participant Characteristics

Of 85 individuals screened, 54 were excluded — the large majority because their eGFR did not fall within the predefined eligibility range — leaving 31 who were enrolled. Of these, 4 discontinued because they no longer wished to continue the study (none for adverse events or product-related reasons), and 27 completed the 90-day intervention (completion rate 87%). Because all 4 discontinuations were due to voluntary withdrawal unrelated to treatment or tolerability, the risk of efficacy-related attrition bias is considered low. Mean age of completers was 53.1 ± 9.5 years (range 38–71); 17 men and 10 women. The participant flow is shown in Figure 1, and baseline characteristics are summarized in Table 1.

**Figure 1.**
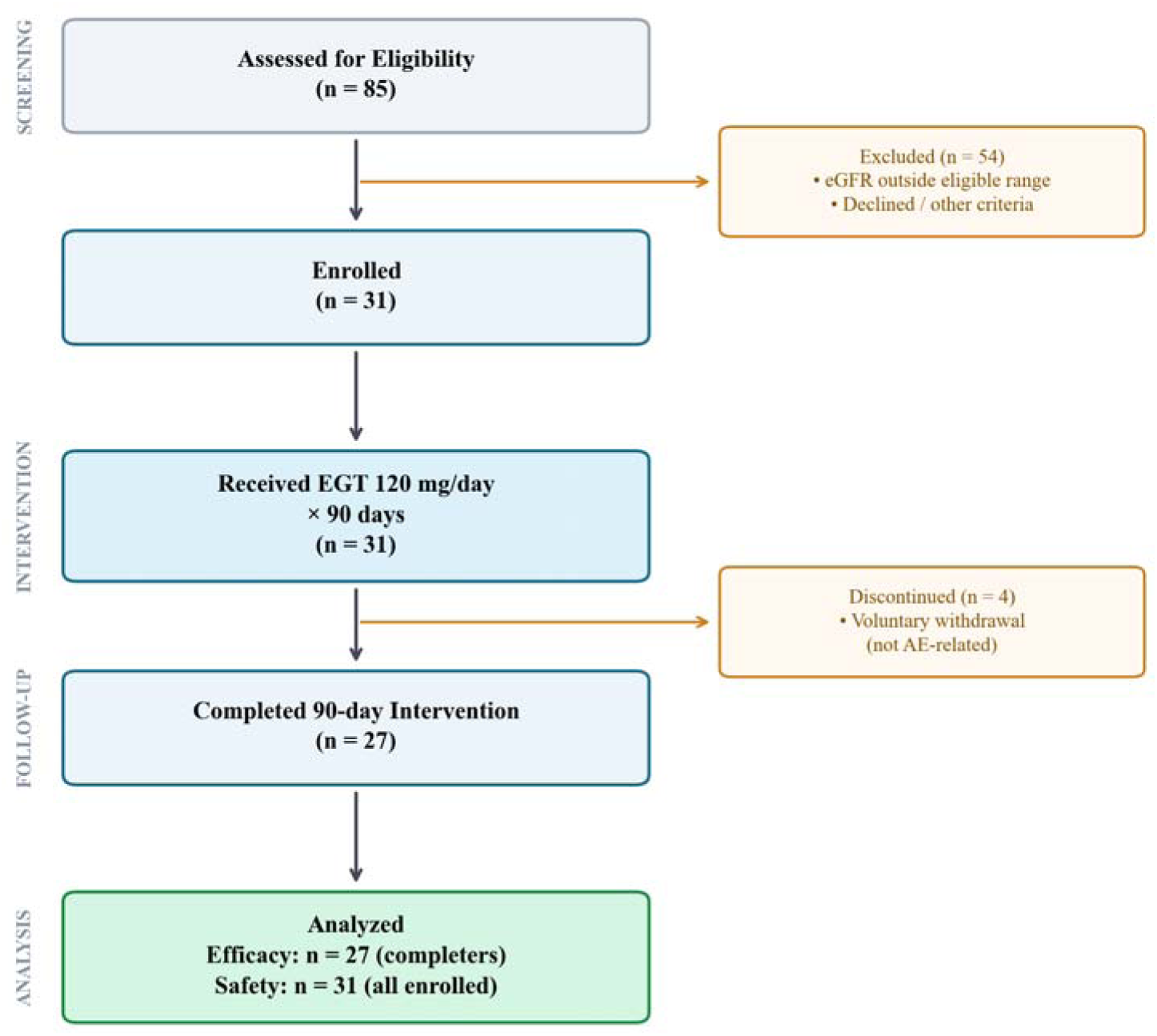
Participant flow diagram (single-arm). 85 screened, 31 enrolled, 27 completed the 90-day intervention.

**Table 1.** Baseline characteristics (N = 27).

| Characteristic | Value |
| --- | --- |
| Age (years) | 53.1 ± 9.5 |
| Sex (male / female) | 17 / 10 |
| BMI (kg/m<sup>2</sup>) | 24.8 ± 2.1 |
| eGFR-creatinine (mL/min/1.73 m<sup>2</sup>) | 86.04 ± 17.89 |
| eGFR-cystatin C (mL/min/1.73 m<sup>2</sup>) | 79.53 ± 21.83 |
| Serum creatinine (μmol/L) | 85.56 ± 23.94 |
| Cystatin C (mg/L) | 1.16 ± 0.36 |
| UACR (mg/g) | 78.84 ± 88.28 |
| Voiding diary total score | 13.67 ± 6.19 |
| Low-back-pain VAS | 2.26 ± 1.75 |

**Table 2.** Within-subject changes in renal function over 90 days (N = 27).

| Outcome | Baseline (D0) | Day 60 | Day 90 | Change (D90–D0) | p (D90 vs D0) |
| --- | --- | --- | --- | --- | --- |
| eGFR-creatinine (mL/min/1.73 m<sup>2</sup>) | 86.04 ± 17.89 | 90.31 ± 20.03 | 93.25 ± 19.00 | +7.20 ± 11.69 (+8.4%) | 0.0016 |
| eGFR-cystatin C (mL/min/1.73 m<sup>2</sup>) | 79.53 ± 21.83 | 83.16 ± 25.10 | 83.13 ± 22.89 | +3.60 ± 12.57 | 0.31 (ns) |
| Serum creatinine (μmol/L) | 85.56 ± 23.94 | 82.11 ± 24.34 | 79.60 ± 22.79 | −5.97 ± 11.76 (−7.0%) | 0.015 |
| Serum urea (mmol/L) | 6.80 ± 1.73 | 6.13 ± 1.55 | 6.29 ± 1.77 | −0.51 ± 1.89 | 0.18 (ns) |
| Cystatin C (mg/L) | 1.16 ± 0.36 | 1.14 ± 0.39 | 1.16 ± 0.39 | 0.00 ± 0.16 | 0.99 (ns) |
| Uric acid (μmol/L) | 331.3 ± 99.9 | 348.8 ± 131.3 | 327.5 ± 91.6 | −3.85 | 0.10 (ns) |
| UACR (mg/g) | 78.84 ± 88.28 | 59.13 ± 68.98 | 67.58 ± 89.24 | −11.26 ± 85.82 (−14.3%) | 0.67 (ns) |

**Table 3.** Within-subject changes in voiding diary and low-back-pain VAS (N = 27).

| Measure | Baseline (D0) | Day 90 | % change | p |
| --- | --- | --- | --- | --- |
| Voiding diary total score | 13.67 ± 6.19 | 5.85 ± 3.69 | -57.2% | <0.0001 |
| Low-back-pain VAS | 2.26 ± 1.75 | 0.74 ± 0.66 | -67.2% | 0.0002 |
| Sensation of incomplete emptying (Q1) | 1.04 | 0.22 | -78.6% | 0.001 |
| Urinary frequency (Q2) | 1.04 | 0.41 | -60.7% | 0.026 |
| Urgency (Q4) | 0.85 | 0.22 | -73.9% | 0.009 |
| Weak stream (Q5) | 1.04 | 0.11 | -89.3% | 0.0002 |
| Straining to void (Q6) | 0.96 | 0.11 | -88.5% | 0.004 |
| Increased urinary foam (Q8) | 2.67 | 1.48 | -44.4% | 0.013 |
| Low-back soreness (Q10) | 1.96 | 0.78 | -60.4% | 0.003 |
| Nocturia (Q9) | 1.74 | 1.30 | -25.5% | 0.08 (ns) |

### 3.2 Renal Function (Primary and Secondary)

Creatinine-based eGFR improved progressively and significantly (+8.4% at Day 90; p = 0.0016), accompanied by a significant decline in serum creatinine (−7.0%; p = 0.015) (Figure 2A, 2C). Notably, cystatin C-based eGFR and serum cystatin C did not change significantly (Figure 2B), indicating that the improvement seen with the creatinine-based equation was not corroborated by an independent, muscle-mass-independent filtration marker. UACR and urea showed non-significant downward trends, and urinary electrolytes (Na, K, Cl) remained stable throughout (all p > 0.05). The distribution of individual eGFR responses is shown in Figure 4, and a summary of within-subject percentage changes across all outcomes is provided in Figure 5.

**Figure 2.**
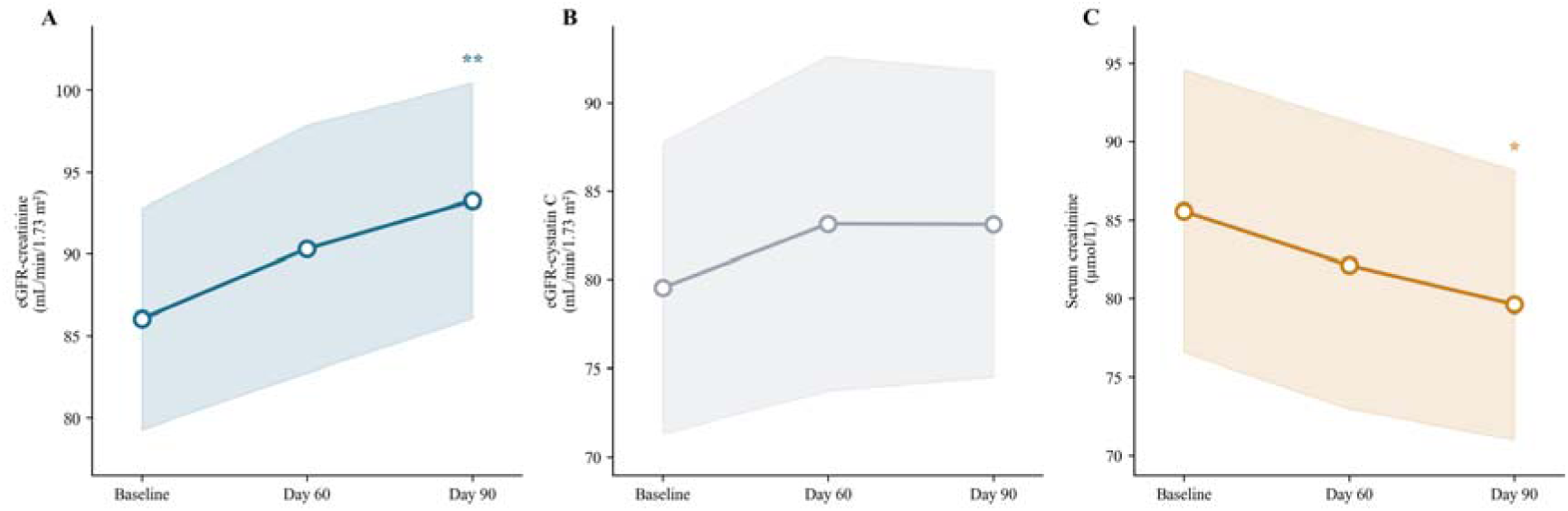
Within-subject changes in renal function over 90 days (N = 27). (A) Creatinine-based eGFR (mL/min/1.73 m²); (B) Cystatin C-based eGFR (mL/min/1.73 m²); (C) Serum creatinine (µmol/L). Mean ± 95% CI; symbols above each timepoint denote paired comparison vs baseline (* p<0.05, ** p<0.01, *** p<0.001). Creatinine-based eGFR improved significantly while cystatin C-based eGFR did not.

**Figure 3.**
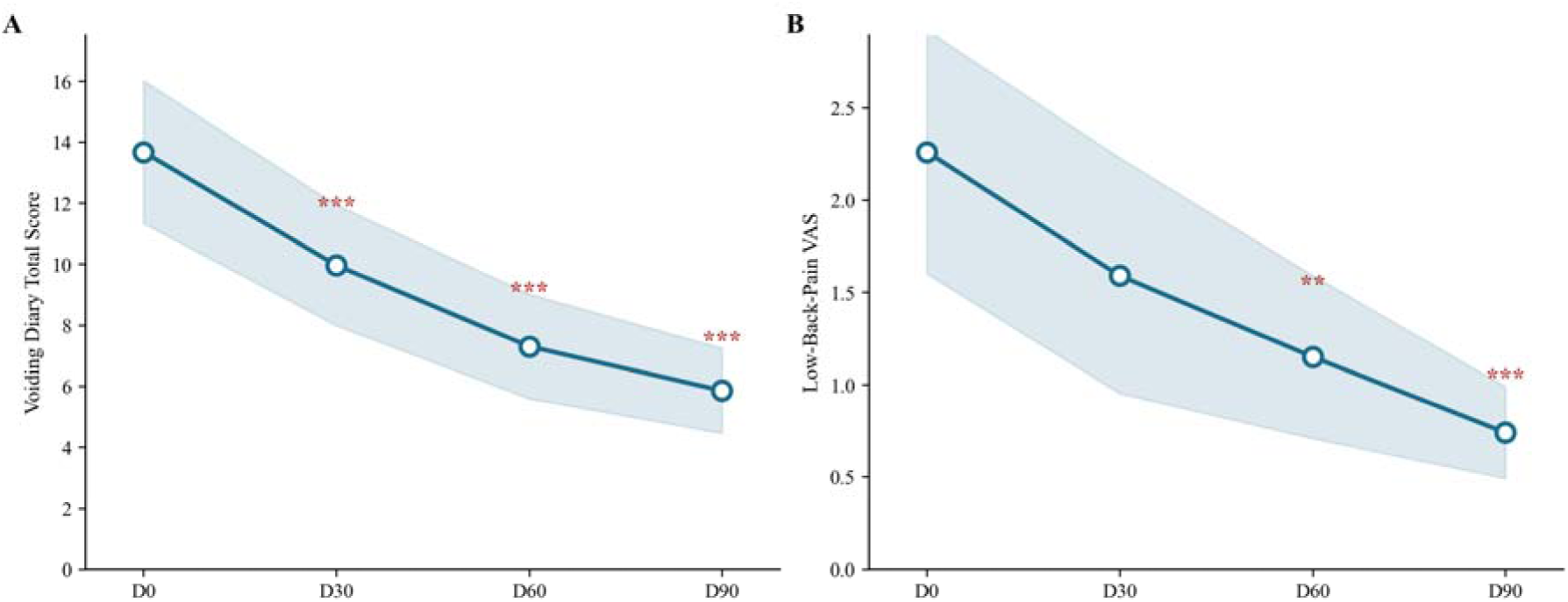
Time-course of patient-reported symptoms (N = 27). (A) Voiding diary total score; (B) Low-back-pain VAS. Mean ± 95% CI; symbols above each timepoint denote paired comparison vs baseline (* p<0.05, ** p<0.01, *** p<0.001).

**Figure 4.**
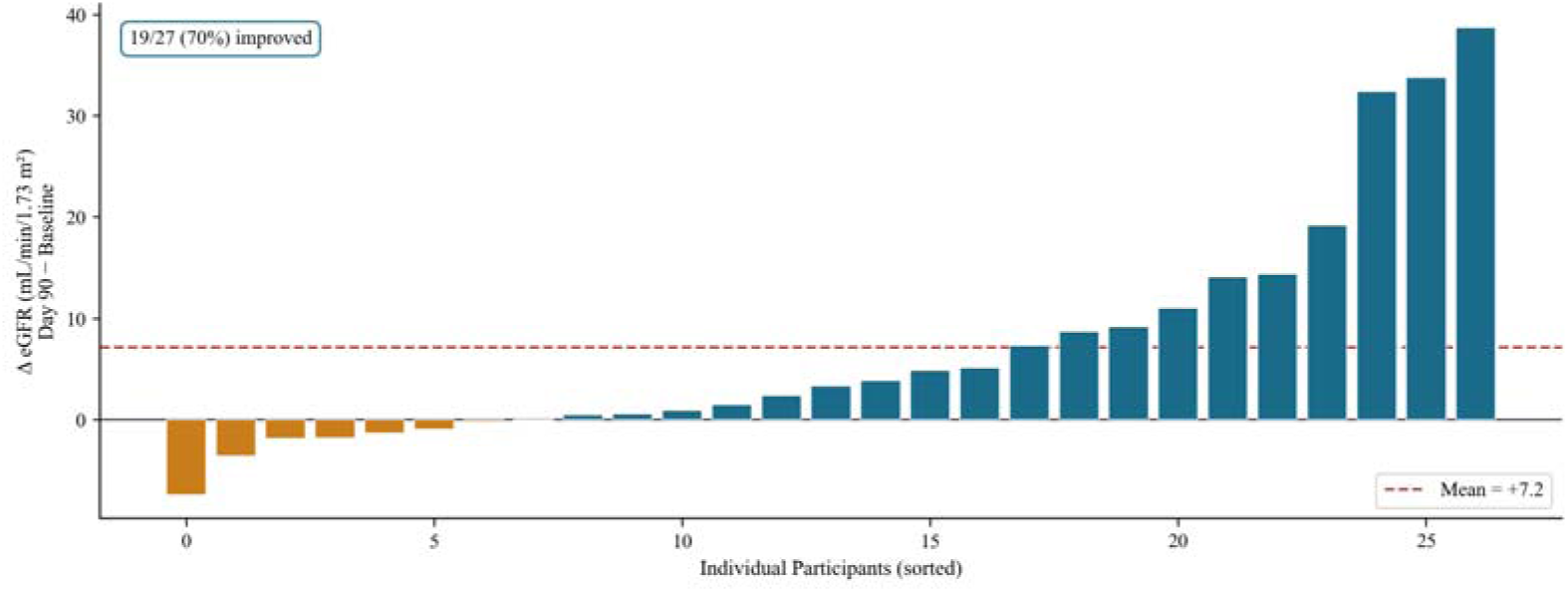
Individual changes in creatinine-based eGFR at Day 90 (N = 27). Per-participant within-subject change. Uncontrolled before-after data.

**Figure 5.**
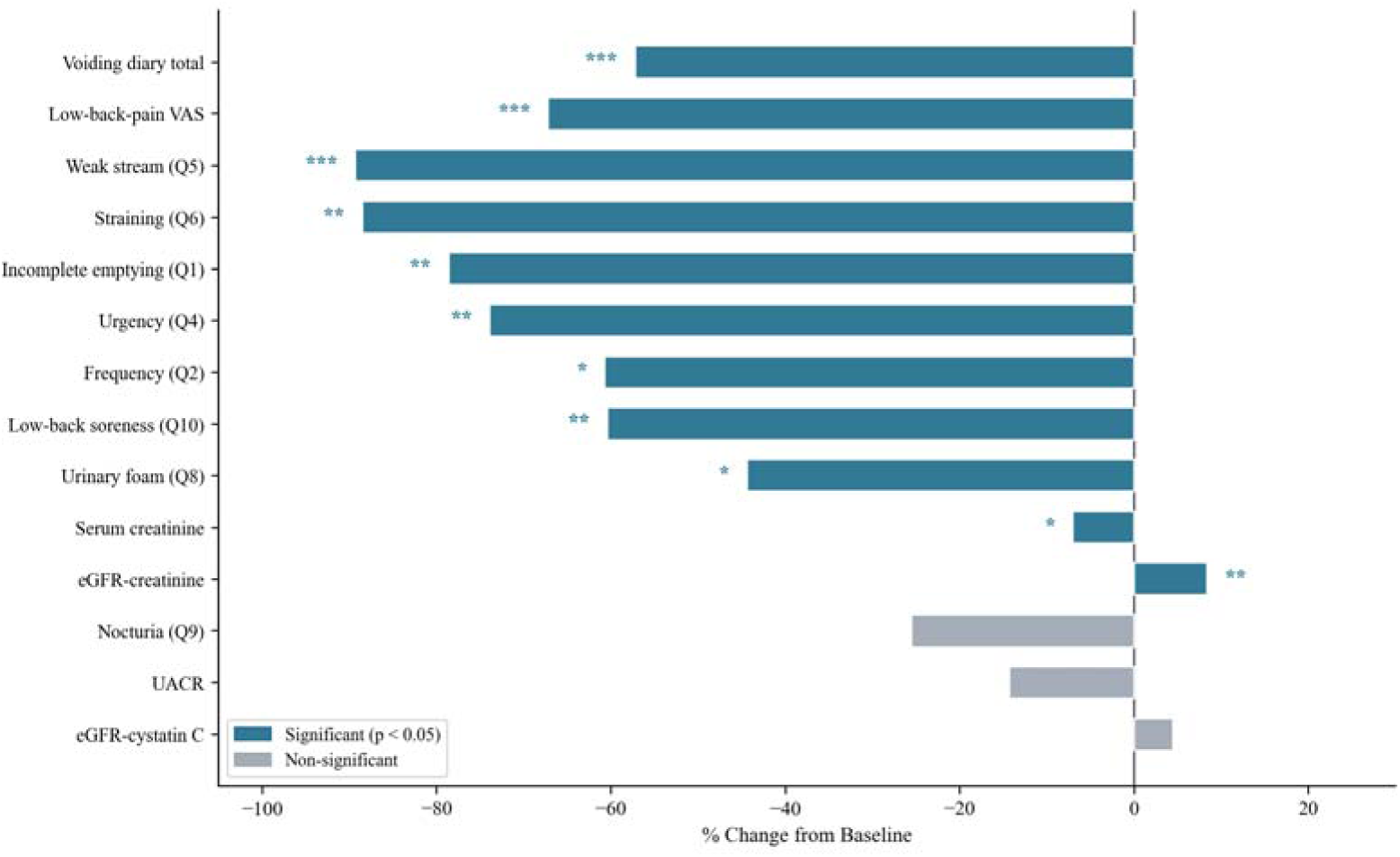
Within-subject percentage change across outcomes at Day 90 (N = 27). Symbols denote paired comparison vs baseline (* p<0.05, ** p<0.01, *** p<0.001).

### 3.3 Patient-Reported Urological Symptoms

Patient-reported symptoms showed the most robust and consistent improvement. The total voiding diary score decreased by 57.2% (p < 0.0001) and low-back-pain VAS by 67.2% (p = 0.0002), both improving progressively across visits (D30 → D60 → D90) (Figure 3A, 3B). Nine of ten diary items improved significantly, with the largest reductions in weak stream, straining to void, and incomplete emptying. Notably, the score for increased urinary foam — a subjective indicator that may parallel glomerular protein leakage — decreased by 44.4% (p = 0.013), directionally consistent with the downward UACR trend. Only nocturia (Q9) did not reach statistical significance.

### 3.4 Exploratory Subgroup Analysis

When stratified by baseline albuminuria, the improvement in creatinine-based eGFR was greater in participants with normal albuminuria (UACR ≤ 30 mg/g: +10.88 ± 14.31, p = 0.014; n = 14) than in those with elevated albuminuria (UACR > 30 mg/g: +3.25 ± 6.46, p = 0.095; n = 13) (Figure 6). The between-subgroup difference did not reach significance (p = 0.09). This exploratory observation suggests that the eGFR signal was driven more by participants with preserved baseline filtration, consistent with regression toward the mean and warranting cautious interpretation.

**Figure 6.**
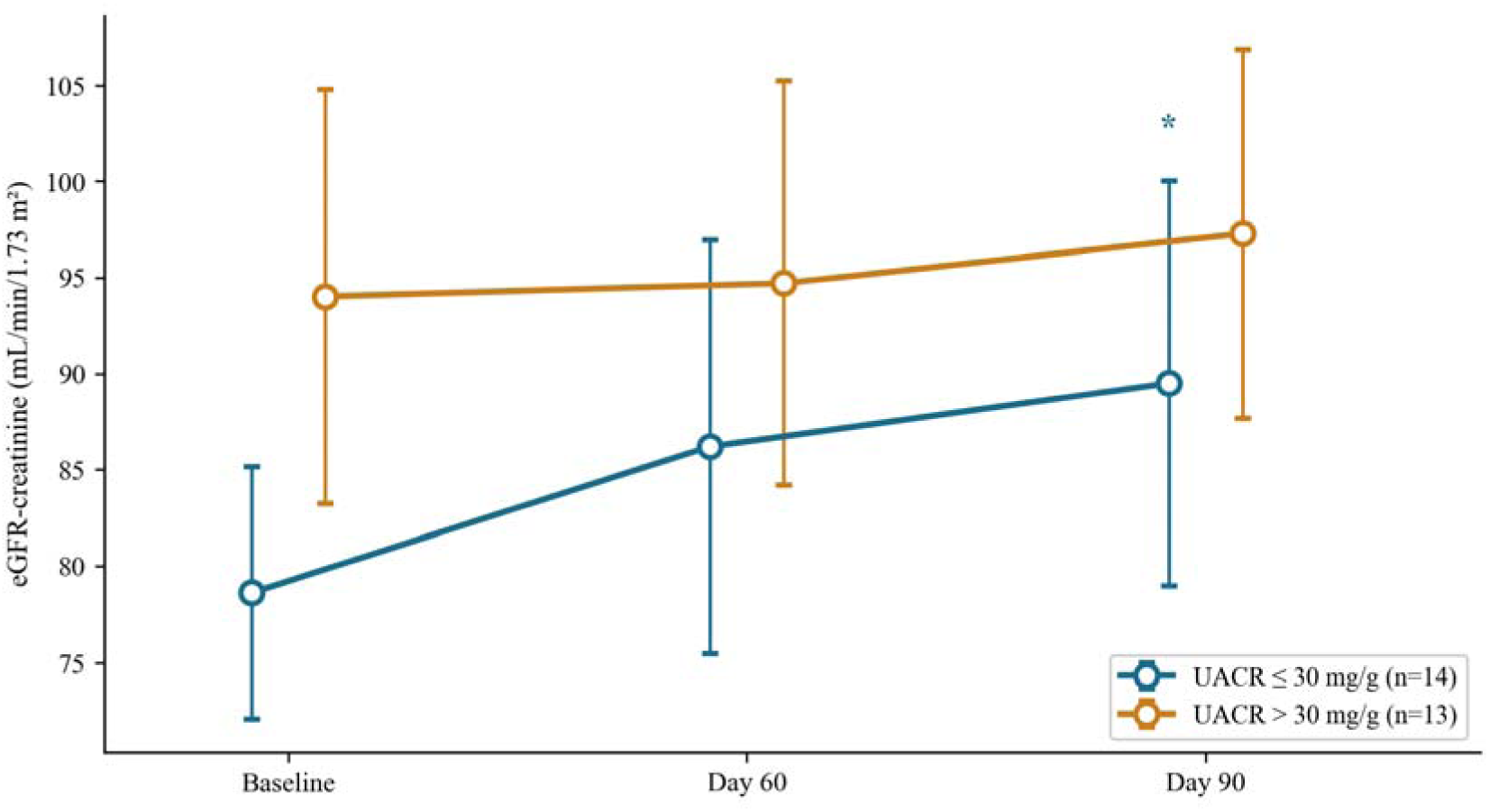
eGFR response stratified by baseline albuminuria (exploratory). Groups offset horizontally for clarity. Symbols at Day 90 = paired comparison vs baseline within subgroup. Between-subgroup difference not significant (p = 0.09).

### 3.5 Systemic Oxidative Stress (Exploratory)

As an exploratory marker of systemic oxidative DNA damage, urinary 8-hydroxy-2’-deoxyguanosine (8-OHdG, normalized to urinary creatinine) was measured at baseline, Day 60, and Day 90. Contrary to the expectation of a systemic antioxidant effect, 8-OHdG did not decrease; rather, it increased from 142.2 ± 59.9 to 169.9 ± 64.8 ng/mg Cr (+27.8; p = 0.044) over 90 days (Figure 7). This finding indicates that the renal-functional and symptomatic changes observed in this cohort were not accompanied by a measurable reduction in systemic oxidative DNA damage. Given the uncontrolled design and the known intra-individual variability of urinary 8-OHdG, this exploratory result should be interpreted with considerable caution; it nonetheless argues against a generalized systemic antioxidant mechanism and is consistent with EGT’s proposed action being tissue-localized rather than systemic.

**Figure 7.**
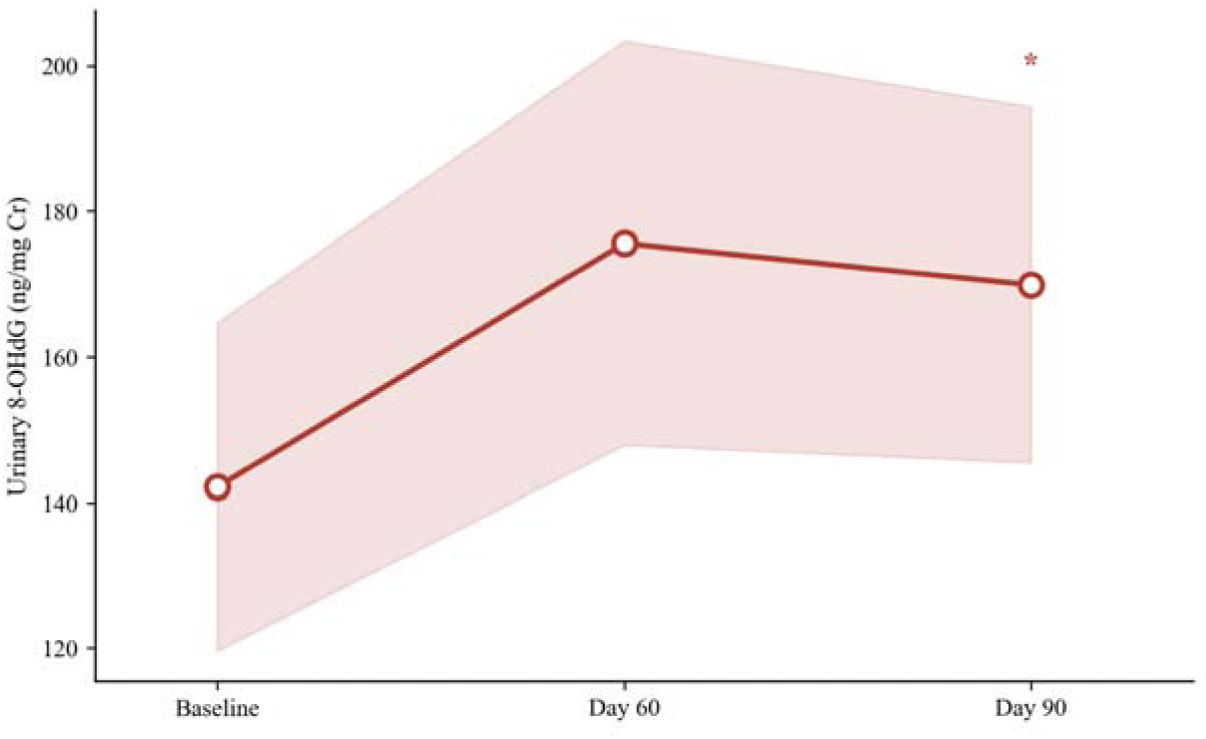
Urinary 8-OHdG over 90 days (exploratory). Systemic oxidative-DNA-damage marker rose rather than fell (* p<0.05 vs baseline), arguing against a systemic antioxidant effect.

### 3.6 Safety

Among the 31 participants in the safety analysis, no product-related adverse events were observed over 90 days. Routine hematology, liver function, and urinalysis showed no clinically significant changes, confirming good tolerability of EGT at 120 mg/day.

## 4. Discussion

This single-center, open-label, self-controlled trial found that 90-day EGT supplementation in adults with early renal function decline was associated with two principal observations: a marked, progressive improvement in patient-reported urological symptoms and quality of life, and an improvement in creatinine-based eGFR. Because the study lacked a control group, these changes cannot be causally attributed to EGT and may partly reflect natural fluctuation, regression to the mean, or placebo effects — a consideration that is especially relevant for the subjective symptom outcomes.

The symptom findings were the most robust. The 57% reduction in total voiding diary score and 67% reduction in low-back-pain VAS (both p < 0.001), improving progressively over the intervention period, represent clinically meaningful relief of the bothersome symptoms that most affect quality of life in this population [4,5]. However, open-label symptom self-report is particularly susceptible to expectation and placebo effects, and these results require confirmation under blinded, controlled conditions.

The filtration findings warrant caution. Although creatinine-based eGFR rose significantly (+8.4%), this was not corroborated by cystatin C-based eGFR or serum cystatin C, both of which remained essentially unchanged. Because serum creatinine is influenced by non-renal factors — including dietary protein, muscle mass, and hydration — the discordance between the two filtration markers raises the possibility that the creatinine-based signal reflects, at least in part, changes in creatinine generation or handling rather than a true increase in glomerular filtration rate. Cystatin C, being largely independent of muscle mass, did not support a genuine filtration gain. This marker discordance is an important limitation and tempers any claim of renoprotection. Future controlled studies should incorporate measured GFR or paired creatinine-cystatin C equations to resolve this question.

The mechanistic rationale for EGT in renal tissue is biologically plausible: OCTN1-mediated accumulation of EGT in proximal tubular cells could support local mitochondrial protection [15,16,18]. Of particular interest, the exploratory measurement of urinary 8-OHdG — a marker of systemic oxidative DNA damage — showed no reduction and indeed a modest increase over the intervention period (Figure 7). While this exploratory result must be interpreted cautiously given the uncontrolled design and the marker’s known variability, it argues against a generalized systemic antioxidant effect and is at least consistent with the hypothesis that any genuine action of EGT is tissue-localized — concentrated within renal proximal tubular cells via OCTN1 — rather than reflected in circulating or systemic oxidative indices. Nonetheless, the present uncontrolled design does not allow us to confirm this mechanism, and no intra-renal oxidative endpoints were available to substantiate it directly.

Relationship to prior EGT research. Our observations should be interpreted against the existing, predominantly preclinical, EGT literature. Salama et al. demonstrated that EGT mitigates cisplatin-induced nephrotoxicity in rodents by activating Nrf2 and suppressing NF-κB and apoptotic signaling [13], while Peng et al. used proteomic analysis to show that EGT attenuates the acute kidney injury–to–chronic kidney disease transition, partly through enhanced antioxidant activity and complement regulation [15]. These animal studies provide a coherent mechanistic basis for a renoprotective effect, but they were conducted in models of acute, chemically induced injury rather than the slow, early-stage functional decline studied here. The clinical relevance of EGT to renal physiology is further underscored by Meyer et al., who found that EGT is removed by hemodialysis far in excess of the amount normally excreted by the kidney, implying that the kidney actively conserves this antioxidant and that its depletion may accompany advancing renal disease [16]. Our finding of symptomatic and creatinine-based improvement in a free-living human cohort is directionally consistent with this body of work, yet it also exposes a key gap: the absence, until now, of controlled human data linking EGT supplementation to validated renal endpoints. The discordance we observed between creatinine- and cystatin C-based eGFR has not, to our knowledge, been examined in prior EGT studies and represents an important consideration for the field — particularly because EGT, as an amino acid derivative that may influence muscle metabolism, could in principle affect creatinine generation independently of true filtration.

Clinical and translational implications. If the symptomatic benefits observed here can be confirmed under controlled conditions, EGT could occupy a distinctive niche: a well-tolerated, orally available nutritional adjunct for the large population with early renal functional decline and bothersome urological symptoms who currently have few options beyond risk-factor control. The progressive, time-dependent pattern of symptom improvement across visits (D30 → D60 → D90) is at least consistent with a cumulative biological effect, although it is equally compatible with a sustained placebo response. Importantly, the stability of urinary electrolytes and the absence of any adverse changes in hepatic or hematologic safety parameters indicate that any such benefit would not come at the cost of disturbing tubular electrolyte handling or systemic safety. These considerations position EGT as a rational candidate for a definitive randomized trial rather than as an established therapy.

Limitations. Several limitations must be emphasized. First and most importantly, the single-arm, open-label design without a control group precludes any causal inference. This concern is most acute for the patient-reported outcomes: because participants were aware they were receiving an active supplement, the marked symptom improvements are particularly vulnerable to placebo, expectation, and Hawthorne effects, as well as to the natural fluctuation of symptoms over time. The magnitude of symptom change observed here should therefore not be assumed to reflect a true pharmacological effect of EGT until confirmed under blinded, placebo-controlled conditions. Second, the urological symptom diary was a study-specific instrument developed in-house and was not subjected to formal psychometric validation (reliability, validity, or responsiveness testing); its scores should be interpreted as exploratory, and future studies should employ validated instruments such as the International Prostate Symptom Score (IPSS) or the Overactive Bladder Questionnaire (OAB-q). Third, the discordance between creatinine- and cystatin C-based eGFR undermines any confident claim of a true improvement in glomerular filtration, as discussed above. Fourth, the study population, although meeting the eligibility criteria for early renal function decline, had a mean baseline eGFR (≈86 mL/min/1.73 m²) close to the normal range; the eGFR improvements observed cannot be distinguished from regression toward the mean, and this cohort is better characterized as having early or borderline renal functional decline rather than established CKD. Fifth, the sample size (N = 27 completers) is small and the study was conducted at a single center, limiting generalizability. Sixth, the 90-day duration is too short to assess hard renal endpoints such as progression to end-stage renal disease. Collectively, these limitations mean the present findings should be regarded as hypothesis-generating only.

## 5. Conclusions

In this single-arm, open-label, self-controlled trial, 90-day supplementation with EGT (120 mg/day) in adults with early renal function decline was associated with marked improvement in patient-reported urological symptoms and low-back pain, and with an improvement in creatinine-based eGFR that was not confirmed by cystatin C. EGT was well tolerated with no product-related adverse events. Owing to the absence of a control group and the discordance between filtration markers, these results cannot establish efficacy and should be interpreted as preliminary. A randomized, double-blind, placebo-controlled trial — ideally incorporating a validated symptom instrument and muscle-mass-independent filtration markers — is warranted to confirm these observations.

## Clinical Trial Registration

This trial was registered with the Chinese Clinical Trial Registry (Registration Number: ChiCTR2500108897) on September 08, 2025 (Prospectively registered).

## Data Availability Statement

The datasets generated and/or analyzed during the current study are not publicly available due to participant privacy but are available from the corresponding author upon reasonable request.

## Authors’ Contributions

Rong Fan: Investigation, Project administration, Supervision, Data curation, and Writing – review & editing.

Zhili Wu: Investigation, Data curation, Formal analysis, and Writing – review & editing.

Yuanyuan Xu: Investigation, Data curation, Formal analysis, and Writing – review & editing.

Wei Liu: Formal analysis, Visualization, Writing – original draft, and Writing – review & editing.

Guohan Zhou: Methodology, Formal analysis, Project administration, and Writing – review & editing.

Wei Ding: Data curation, Formal analysis, and Writing – review & editing.

Juan Cao: Data curation, Formal analysis, and Writing – review & editing.

Guohua Xiao: Supervision, Funding acquisition, Project administration, Formal analysis, and Writing – review & editing.

Dalin Xu: Investigation, Project administration, Clinical supervision, and Writing – review & editing.

Huan Zhou: Conceptualization, Methodology, Investigation, Project administration, Supervision, Interpretation of results, and Writing – review & editing.

Rong Fan, Zhili Wu, Yuanyuan Xu, Wei Liu, and Guohan Zhou contributed equally to this work and share first authorship. Guohua Xiao, Dalin Xu, and Huan Zhou are co-corresponding authors. All authors reviewed and approved the final manuscript and agreed to be accountable for the integrity and accuracy of the work.

## Acknowledgments

The authors sincerely thank all participants who took part in this study. The authors also thank the clinical and research staff of the Clinical Research Hospital of the First Affiliated Hospital of Anhui Medical University and Lujiang County People’s Hospital for their assistance with participant recruitment, clinical assessments, sample collection, and study coordination.

## References

1. KDIGO 2024 Clinical Practice Guideline for the Evaluation and Management of Chronic Kidney Disease (CKD). Kidney Int. 2024. 10.1016/j.kint.2023.10.018.

2. Jha V, Garcia-Garcia G, Iseki K, et al. Chronic kidney disease: global dimension and perspectives. Lancet. 2013;382(9888):260–272. 10.1016/S0140-6736(13)60687-X.

3. Levin A, Tonelli M, Bonventre J, et al. Global kidney health 2017 and beyond: a roadmap for closing gaps in care, research, and policy. Lancet. 2017;390(10105):1888–1917. 10.1016/S0140-6736(17)30788-2.

4. Cavanaugh KL. Patient-reported outcomes in chronic kidney disease: the way forward. Clin J Am Soc Nephrol. 2014;9(7):1273–1281. 10.2215/CJN.10841013.

5. Deshpande PR, Rajan S, Sudeepthi BL, et al. Patient-reported outcomes: a new era in clinical research. Perspect Clin Res. 2011;2(4):137–144. 10.4103/2229-3485.86879.

6. Galvan DL, Green NH, Danesh FR. Mitochondria in the pathogenesis of human kidney diseases. Nat Rev Nephrol. 2017;13(12):749–762. 10.1038/nrneph.2017.135.

7. Jun M, Venkataraman V, Razavian M, et al. Antioxidants for chronic kidney disease. Cochrane Database Syst Rev. 2012;(10):CD008176. 10.1002/14651858.CD008176.pub2.

8. Gründemann D, Harlfinger S, Golz S, et al. Discovery of the ergothioneine transporter. Proc Natl Acad Sci U S A. 2005;102(14):5256–5261. 10.1073/pnas.0408624102.

9. Halliwell B, Cheah IK, Tang RM. Ergothioneine — an underappreciated dietary amino acid with a specific transporter. FEBS Lett. 2018;592(20):3357–3366. 10.1002/1873-3468.13123.

10. Cheah IK, Halliwell B. Ergothioneine; antioxidant potential, physiological function and role in disease. Biochim Biophys Acta. 2012;1822(5):784–793. 10.1016/j.bbadis.2011.09.017.

11. Pochini L, Galluccio M, Scalise M, et al. OCTN cation transporters in health and disease: role as drug targets. Int J Mol Sci. 2013;14(11):21313–21342. 10.3390/ijms141121313.

12. Urban TJ, Gallagher RC, Meyer UA, et al. Functional effects of protein sequence polymorphisms in the organic cation/ergothioneine transporter OCTN1 (SLC22A4). Pharmacogenet Genomics. 2007;17(9):773–782. 10.1097/FPC.0b013e32822a10be.

13. Salama SA, Abd-Allah GM, Mohamadin AM, Elshafey MM, Gad HS. Ergothioneine mitigates cisplatin-evoked nephrotoxicity via targeting Nrf2, NF-κB, and apoptotic signaling and inhibiting γ-glutamyl transpeptidase. Life Sci. 2021; 278: 119572. 10.1016/j.lfs.2021.119572.

14. Mayayo-Vallverdú C, López de Heredia M, Prat E, et al. The antioxidant L-ergothioneine prevents cystine lithiasis in the Slc7a9-/- mouse model of cystinuria. Redox Biol. 2023; 64: 102801. 10.1016/j.redox.2023.102801.

15. Peng J, Chen J, Wu M, et al. Proteomic analysis reveals the potential mechanism of ergothioneine in preventing acute kidney injury to chronic kidney disease transition. Arch Biochem Biophys. 2025; 770: 110534. 10.1016/j.abb.2025.110534.

16. Meyer TW, Aronov PA, Sirich TL, et al. Depletion by hemodialysis of the antioxidant ergothioneine. Kidney360. 2025; 6(2): 288–296. 10.34067/kid.0000000645.

17. Shadel GS, Horvath TL. Mitochondrial ROS signaling in gene expression and aging. Genetics. 2014;198(1):163–187. 10.1534/genetics.114.167387.

18. Smith RA, Murphy MP. Mitochondria-targeted antioxidants in the treatment of kidney disease. Semin Nephrol. 2018;38(2):161–171. 10.1016/j.semnephrol.2018.02.004.

19. Bower WF, et al. Impact of nocturia on health-related quality of life in patients with chronic kidney disease. Nephrology (Carlton). 2020;25(11):854–862. 10.1111/nep.13735.

20. Inker LA, Eneanya ND, Coresh J, et al. New creatinine- and cystatin C-based eGFR equations without race. N Engl J Med. 2021;385(19):1737–1749. 10.1056/NEJMoa2102953.

